# DWI and Clinical Characteristics Correlations in Acute Ischemic Stroke After Thrombolysis

**DOI:** 10.1101/2025.06.03.25328925

**Authors:** Jiayi Li, Cheng Huang, Yangtao Liu, Yingli Li, Jie Zhang, Mi Xiao, Zhiwen Yan, Huan Zhao, XiaoPeng Zeng, Jun Mu

## Abstract

**Objective:** Magnetic Resonance Diffusion-Weighted Imaging (DWI) is a crucial tool for diagnosing acute ischemic stroke, yet some patients present as DWI-negative. This study aims to analyze the imaging differences and associated clinical characteristics in acute ischemic stroke patients receiving intravenous thrombolysis, in order to enhance understanding of DWI-negative strokes.

**Methods:** Retrospective collection of clinical data from acute ischemic stroke patients receiving intravenous thrombolysis at the Stroke Center of the First Affiliated Hospital of Chongqing Medical University from January 2017 to June 2023, categorized into DWI-positive and negative groups. Descriptive statistics, univariate analysis, binary logistic regression, and machine learning model were utilized to assess the predictive value of clinical features. Additionally, telephone follow-up was conducted for DWI-negative patients to record medication compliance, stroke recurrence, and mortality, with Fine-Gray competing risk model used to analyze recurrent risk factors.

**Results:** The incidence rate of DWI-negative ischemic stroke is 22.74%. Factors positively associated with DWI-positive cases include onset to needle time (ONT), onset to first MRI time (OMT), NIHSS score at 1 week of hospitalization (NIHSS-1w), hyperlipidemia (HLP), and atrial fibrillation (AF) (p<0.05, OR>1). Conversely, recurrent ischemic stroke (RIS) and platelet count (PLT) are negatively correlated with DWI-positive cases (p<0.05, OR<1). Trial of Org 10172 in Acute Stroke Treatment (TOAST) classification significantly influences DWI presentation (p=0.01), but the specific impact of etiological subtypes remains unclear. Machine learning models suggest that the features with the highest predictive value, in descending order, are AF, HLP, OMT, ONT, NIHSS difference within 24 hours post-thrombolysis(NIHSS-d(0-24h)PT), and RIS.

**Conclusions:** NIHSS-1w, OMT, ONT, HLP, and AF can predict DWI-positive findings, while platelet count and RIS are associated with DWI-negative cases. AF and HLP demonstrate the highest predictive value. DWI-negative patients have a higher risk of stroke recurrence than mortality in the short term, with a potential correlation between TOAST classification and recurrence risk.

## 1. Introduction

According to the Global Burden of Disease Study 2019, stroke remains the second leading cause of death globally and the third leading cause of combined mortality and disability[1]. Early diagnosis and treatment of stroke lead to better patient outcomes. Ischemic stroke accounts for approximately 62.4% of stroke events worldwide[1]. The main manifestation of acute ischemic stroke is acute-onset focal neurological deficits. The American Neurological Association proposed as early as 2010 that Diffusion-Weighted Imaging (DWI) should be utilized as an effective tool for the early and accurate diagnosis of acute ischemic stroke[1]. The American Heart Association/American Stroke Association’s updated treatment guidelines in 2019 explicitly recommend the use of DWI sequences as an imaging tool to support diagnosis by clinicians[3]. With the widespread application of Diffusion-Weighted Imaging (DWI), an increasing number of studies have identified a subset of patients clinically diagnosed with acute ischemic stroke who do not exhibit high signal intensity on DWI sequences, termed as DWI-negative brain infarction[4, 5]. DWI-negative brain infarction patients account for approximately 6.8%-12.36% of all acute ischemic stroke cases[6–8]. For physicians with limited clinical experience, the presence of DWI-negative findings may lead to underdiagnosis and delayed treatment of acute brain infarction. It is essential to provide alert information for this specific clinical phenomenon to enhance our confidence in diagnosing acute brain infarction. Intravenous thrombolysis is one of the key treatment modalities for patients with acute ischemic stroke, often administered by clinicians prior to DWI scanning. While many researchers have explored the characteristics of DWI-negative brain infarction patients [9], few studies have analyzed the impact of intravenous thrombolysis on the imaging features of patients with acute ischemic stroke. The main objective of this study is to compare the imaging differences and clinical characteristics of patients with acute ischemic stroke after intravenous thrombolysis, aiming to enhance the accurate diagnosis of acute ischemic stroke patients.

## 2. Method

### 2.1 Study Design and Population

In this study, we reviewed the list of patients who underwent intravenous thrombolysis treatment at the Stroke Center of the First Affiliated Hospital of Chongqing Medical University from January 2017 to June 2023. We searched the medical records system to collect information on patients diagnosed with acute ischemic stroke and possessing magnetic resonance imaging data that included DWI sequences. Diagnosis of acute ischemic stroke is based on the sudden onset of focal neurological deficits lasting more than 24 hours or the occurrence of new neurological deficits after a transient ischemic attack, with cranial MRI or CT images during hospitalization showing evidence of acute brain infarction or no significant abnormalities[3]. Finally, two experienced clinicians independently reviewed the imaging studies and reports of the patients, providing diagnoses of DWI-positive and negative brain infarction. Patients were included based on a consensus reached by the two physicians. To minimize the possibility of misdiagnosis, we simultaneously documented the clinical manifestations of acute ischemic stroke in DWI-negative patients. A cerebrovascular disease specialist conducted the diagnosis and differential diagnosis, ultimately retaining patients diagnosed with acute ischemic stroke (Figure 1).

**Figure 1.**
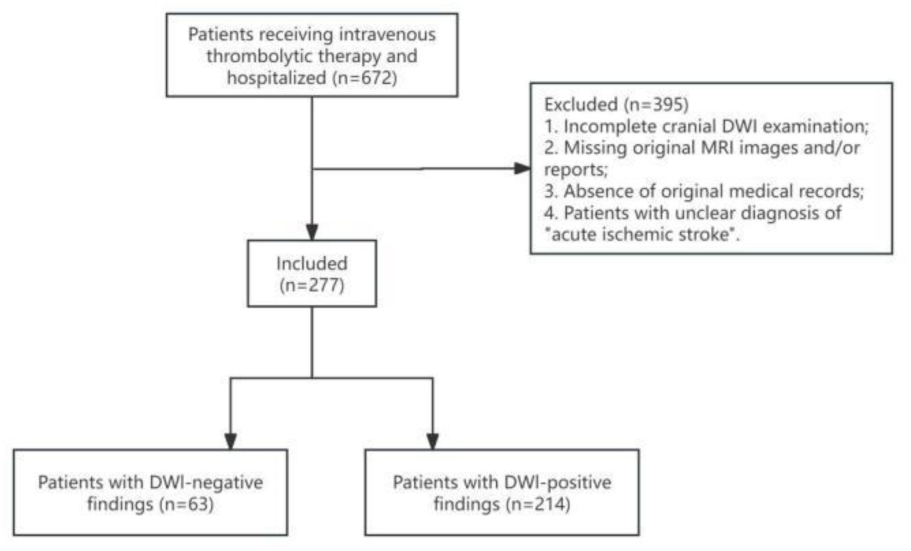
Flowchart of the study population selection. We searched for electronic and paper-based thrombolysis registration data from the Stroke Center of the First Affiliated Hospital of Chongqing Medical University between January 2017 and June 2023. A total of 672 medical records were identified. After screening, we included 277 eligible patients with acute ischemic stroke. Following assessment by two physicians, they were divided into two groups, including 214 DWI-positive ischemic stroke patients and 63 DWI-negative ischemic stroke patients.

The inclusion criteria for this study were as follows: (1) administration of alteplase or urokinase intravenous therapy in the emergency department; (2) immediate hospitalization for inpatient care following intravenous thrombolysis; (3) completion of cranial MRI examination during hospitalization, including DWI sequences; (4) preservation of complete and viewable cranial MRI images; (5) clear diagnosis of acute ischemic stroke in the patient’s medical records and expert opinions. Exclusion criteria for this study were: (1) failure to receive inpatient care after intravenous thrombolysis treatment; (2) incomplete cranial DWI imaging examination; (3) loss of cranial MRI images and reports; (4) inability to access medical records.

Furthermore, to assess the prognosis of DWI-negative brain infarction patients, we conducted telephone follow-up for all patients in the DWI-negative group in November 2023, documenting medication adherence, stroke recurrence, or mortality outcomes.

### 2.2 Data Collection

We documented the DWI findings and collected demographic and baseline data of the enrolled patients, including laboratory indices, cardiovascular and cerebrovascular disease risk factors, four critical durations, and NIHSS scores at four key time points. Risk factors for cardiovascular and cerebrovascular diseases included history of smoking, alcohol consumption, hypertension, diabetes, atrial fibrillation, and hyperlipidemia. The four critical durations referred to the onset to hospital-door time (ODT), the onset to needle time (ONT), the onset to first MRI time (OMT), and the onset to hospital discharge time (OHDT). The NIHSS scores at the four key time points were NIHSS score immediately post-thrombolysis (NIHSS-i PT), NIHSS score at 24 hours post-thrombolysis (NIHSS-24h PT), NIHSS score at 1 week of hospitalization (NIHSS-1w), NIHSS score at hospital discharge (NIHSS-HD).

Additionally, we classified each patient according to TOAST criteria proposed by Adams in 1993[10]. Furthermore, in November 2023, we conducted telephone interviews to collect post-discharge outcomes of DWI-negative brain infarction patients, including recurrence status and time, mortality outcomes and causes, medication discontinuation, and duration of medication adherence.

### 2.3 Statistical Methods

In the descriptive statistical analysis, we presented continuous data using mean ± standard deviation or median with interquartile range, and categorical data using frequencies and percentages. Due to incomplete data in some cases leading to partially random missing data, we conducted a missing value analysis and performed simple imputation using means, medians, and modes. Using IBM SPSS Statistics 25, we conducted normality tests on demographic information, laboratory indices, and clinical characteristics of both the DWI-negative and DWI-positive groups. Independent samples t-tests, chi-square tests, and Mann-Whitney U tests were employed for between-group univariate analysis, with a significance level set at 0.05. Based on the univariate statistical analysis, clinical characteristics showing significant differences were selected as independent variables, with DWI findings as the dependent variable. The Trial of Org 10172 in Acute Stroke Treatment (TOAST) classification was transformed into dummy variables for binary logistic regression analysis.

We utilized Python 3.10.7 to run code and employed regularization techniques of the Support Vector Classification (SVC) model to optimize and select clinical information such as NIHSS scores, medical history, and laboratory indices. This process yielded predictive clinical features. Subsequently, using the scikit-learn 1.2.1 library, we constructed six machine learning models for predicting DWI outcomes: eXtreme Gradient Boosting, Support Vector Machine, Logistic Regression, Random Forest, Gradient Boosting Decision Tree, and Light Gradient Boosting Machine models. We compared the Area Under the Curve (AUC) values of the Receiver Operating Characteristic (ROC) curves of each predictive model to assess predictive performance and selected the model with the best predictive results. Finally, we utilized shap 0.42.1 and matplotlib 3.7.0 for interpreting and visually presenting the results of the optimal model.

Based on telephone interviews regarding stroke recurrence, we categorized DWI-negative patients into recurrent and non-recurrent groups. Prior to the telephone follow-up, some patients had already deceased due to severe cerebral infarction, cancer, myocardial infarction, and other causes, involving multiple competing endpoints. Therefore, we utilized R software to establish a Fine-Gray competing risk model as a survival analysis method to analyze the risk of recurrent ischemic stroke in DWI-negative patients. In the model analysis, patients who did not experience stroke recurrence by the time of the telephone interview and those lost to follow-up were considered as censored data. The time of loss to follow-up or the study start and end times were used as endpoint times. The death of patients due to severe cerebral infarction was considered as a recurrent event. Additionally, the death of patients due to non-cerebral infarction causes such as cancer was regarded as a competing risk event.

### 2.4 Medical Ethics

This retrospective study obtained approval from the Ethics Review Committee of the First Affiliated Hospital of Chongqing Medical University. Due to minimal risk to the participants in this study, the ethics committee granted a waiver of informed consent for the research participants.

## 3. Result

### 3.1 Multilevel Statistical Analysis

From January 2017 to June 2023, a total of 672 patients underwent intravenous thrombolysis treatment at the hospital stroke center. Among them, magnetic resonance imaging data of 277 hospitalized patients were available for complete review. Among these, there were 214 patients with acute ischemic stroke showing DWI-positive findings and 63 patients with DWI-negative acute ischemic stroke. The incidence rate of DWI-negative acute ischemic stroke was 22.74% (63/277). Interestingly, there were 65 cases of DWI-negative stroke patients, with one patient experiencing three episodes of acute ischemic stroke and undergoing three sessions of intravenous thrombolysis, all resulting in negative DWI scans. Among the 63 DWI-negative stroke patients, 8 were lost to follow-up due to incorrect contact information, and 9 died. Among the deceased, 3 died from severe cerebral infarction, while 6 passed away after discharge due to causes such as cancer, myocardial infarction, heart failure, and Coronavirus Disease 2019.

A small amount of clinical data was completely randomly missing, with missing value analysis indicating that 81% of the missing data had a missing rate of less than 10%, and 68% had a missing rate of no more than 5%. After simple imputation of missing values, intergroup univariate analysis of various characteristics between the DWI-negative and DWI-positive groups was conducted (Table 1). It was found that there were no statistically significant differences in age (p=0.540) and gender (p=0.077) between the two groups. However, significant statistical differences were observed in the NIHSS scores at four important time points (P<0.01) and four critical durations (P<0.01). Additionally, the presence of HLP (P=0.001), AF (P=0.000), and RIS (P=0.031) also showed significant statistical differences. To ensure the practical clinical significance of predictive indicators, we introduced a new index, NIHSS-d(0-24h)PT, which represents the difference between the NIHSS score immediately post-thrombolysis and the NIHSS score at 24 hours post-thrombolysis. This index reflects the thrombolytic effect on patients, with a larger value indicating a potentially better thrombolytic effect. During non-parametric testing, we found no statistical difference in NIHSS-d(0-24h)PT (p=0.134) between the DWI-positive and negative groups. To adjust for confounding factors, we further conducted binary logistic regression analysis, using clinical features that showed intergroup statistical differences in the univariate analysis as independent variables. Ultimately, we discovered that the presence of OMT (p=0.020, OR=1.01), ONT (p=0.004, OR=1.01), NIHSS-1w (p=0.001, OR=1.32), HLP (p=0.000,

**Table 1.**
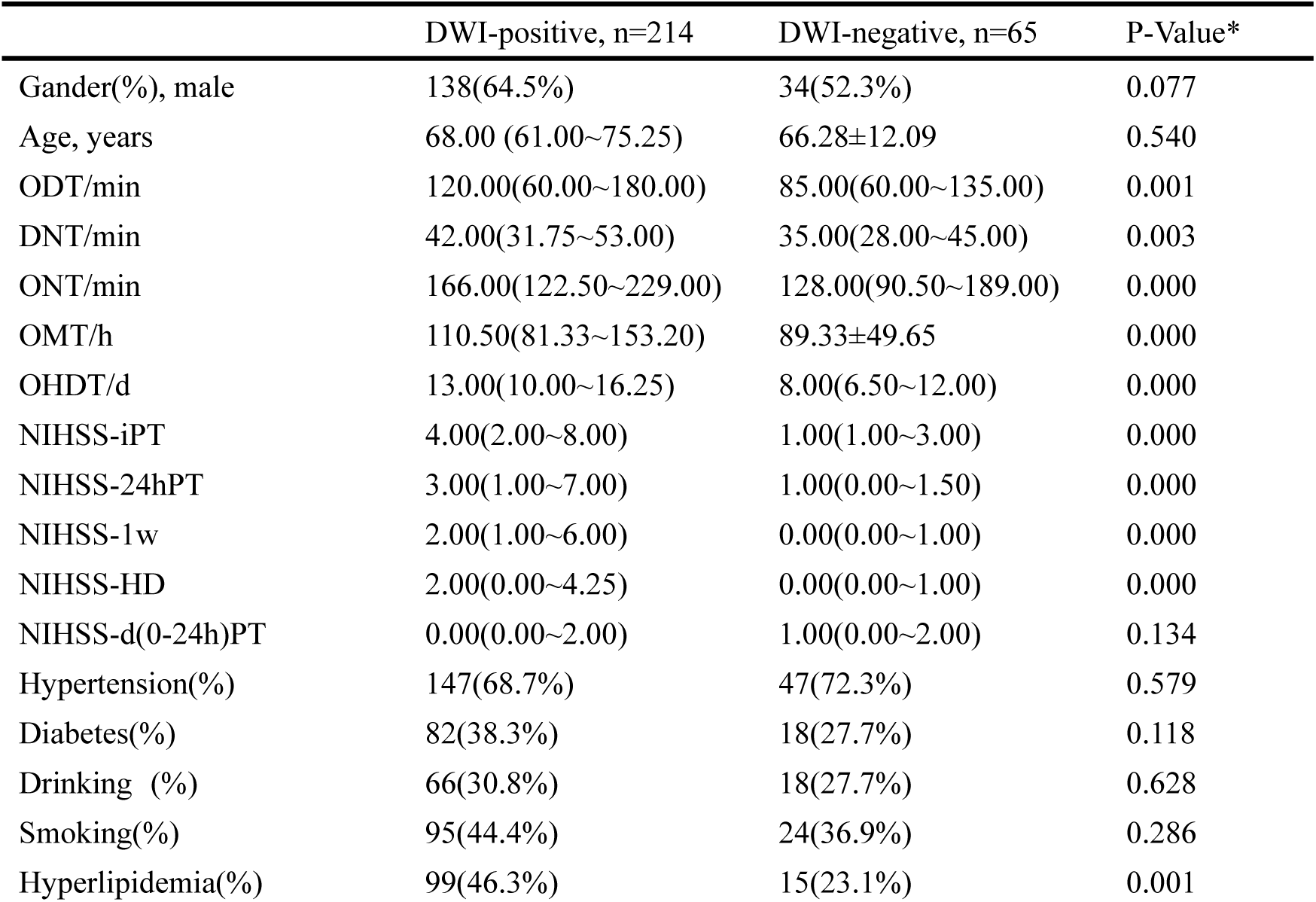
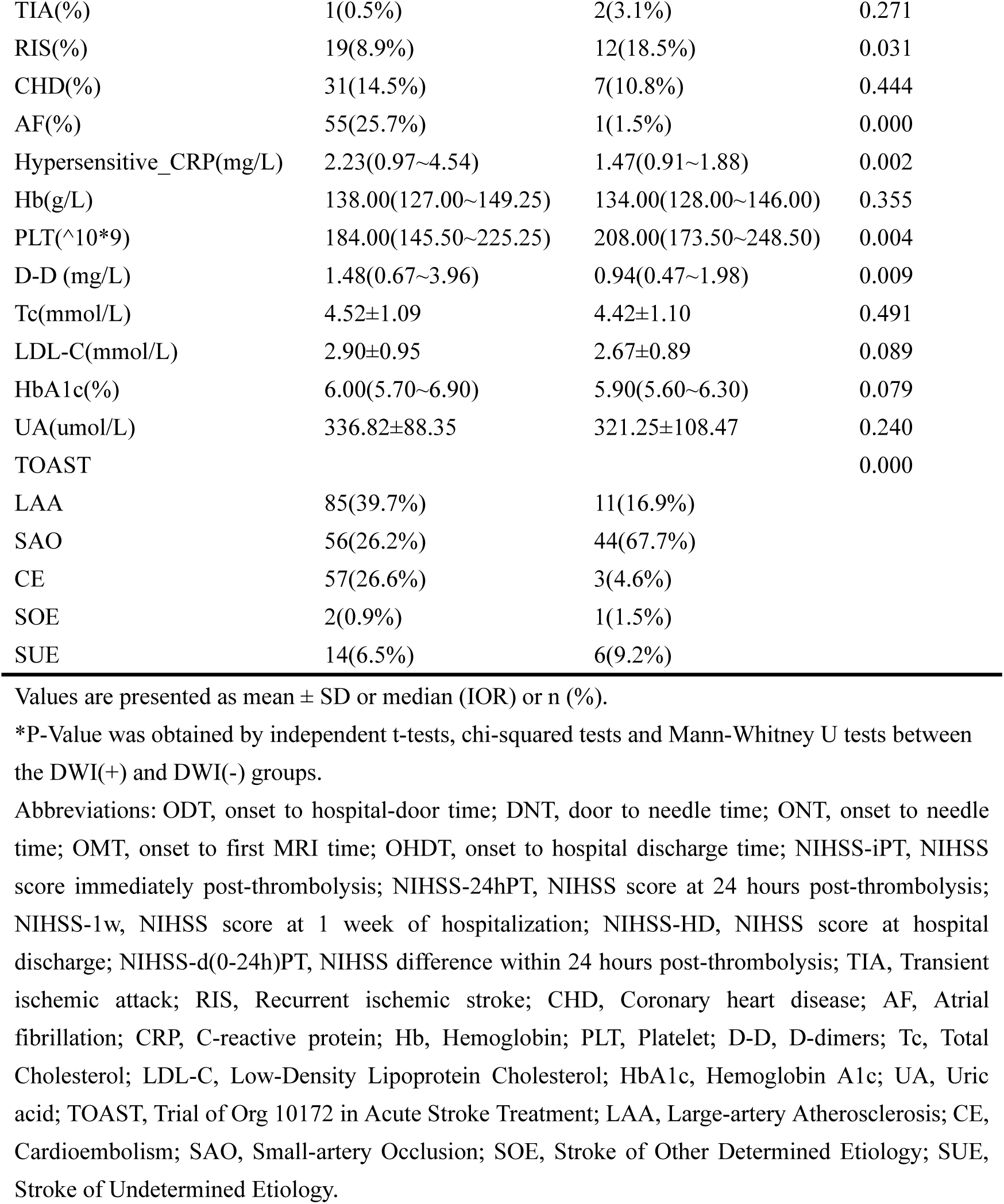
Demographic and clinical characteristics of the study participants.

OR=4.32), and AF (p=0.016, OR=35.94) were positively associated with the likelihood of DWI-positive lesions, while RIS (p=0.037, OR=0.30), PLT (p=0.016, OR=0.99), and the TOAST classification (p=0.011) were negatively associated with the likelihood of DWI-positive lesions. The TOAST classification significantly influenced DWI lesion presentation, although specific categories did not show significant differences compared to large artery atherosclerosis subtype (Table 2). All patients included in this study were diagnosed with acute ischemic stroke. In logistic regression analysis, the dependent variables (DWI-positive and negative) were complementary, suggesting a positive correlation between RIS, PLT, and the likelihood of DWI-negative lesions. In summary, the results of the logistic analysis indicate that NIHSS-1w, OMT, ONT, PLT, HLP, AF, RIS, and TOAST classification are independent predictive factors for DWI-positive lesions. The comparison of the remaining TOAST subtypes with large artery atherosclerosis subtype did not show statistical significance (p>0.05), indicating that the specific type of stroke etiology requires further investigation regarding its impact on DWI presentation.

**Table 2.**
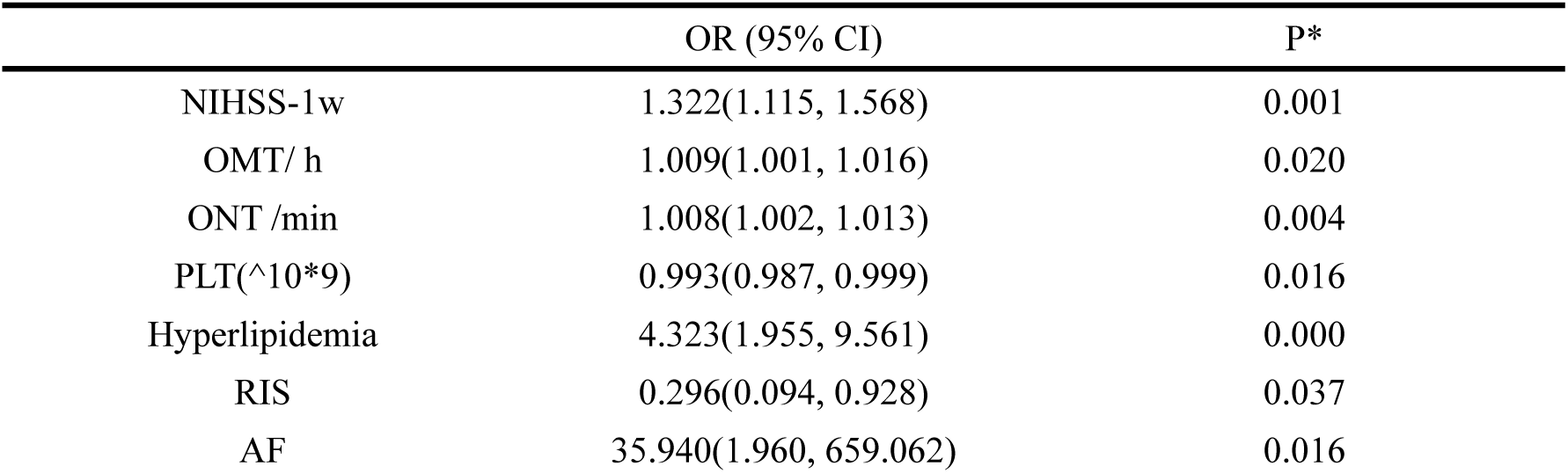
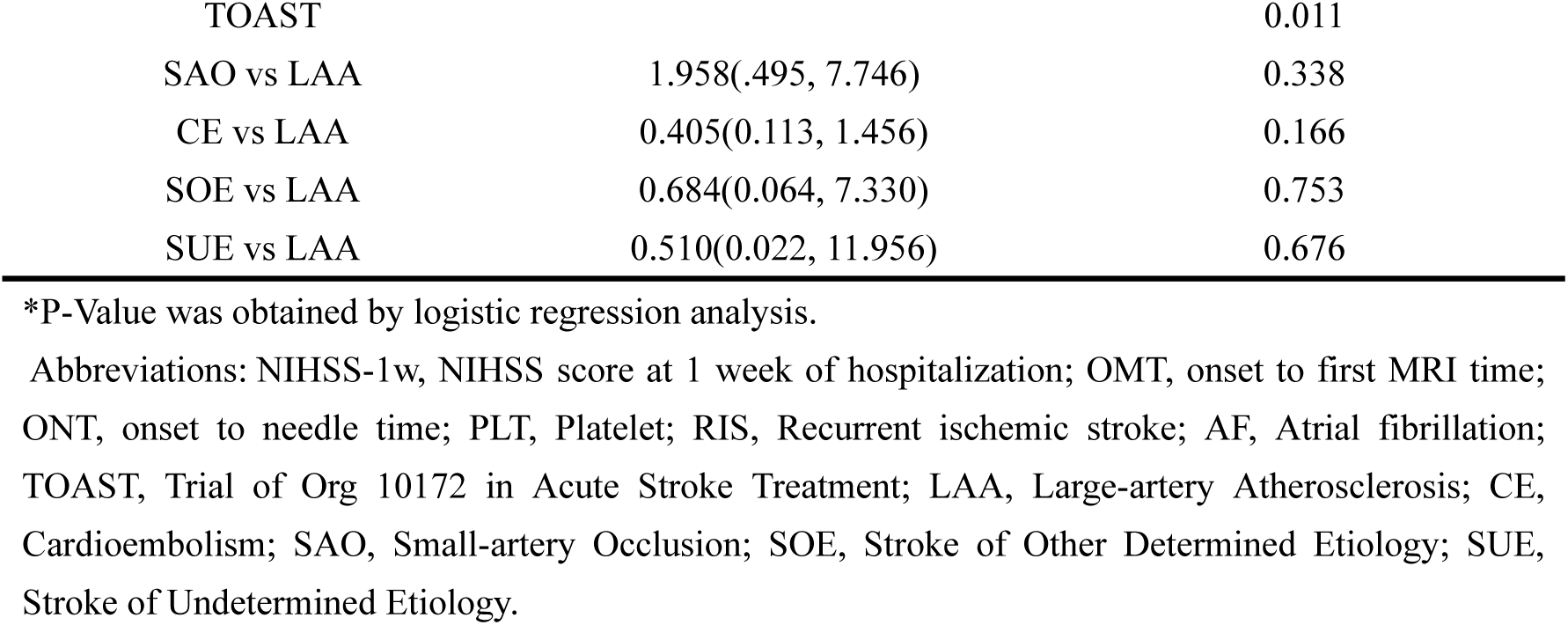
Association between clinical manifestations and DWI lesions.

### 3.2 Machine Learning Models

After regularization processing of the SVC model, six clinically relevant features were identified for prediction: AF, HLP, RIS, ONT, OMT, and NIHSS-d(0-24h)PT. The variance inflation factor for all these clinical features was less than 10, indicating weak collinearity among the aforementioned features. We evaluated the predictive performance of six models by comparing the AUC values of their ROC curves (Figure 2.A). The results indicated that the Logistic Regression (LR) model performed the best, with an AUC value of 0.825. The precision and recall rates for the test set were 0.930 and 0.727, respectively, suggesting that this model has good predictive efficacy and is valuable in minimizing false positives. In this model, a higher average absolute value of the SHAP values for clinical features indicates a stronger impact on the predictive probability. A history of atrial fibrillation is the most significant predictive indicator for DWI-positive findings, exerting a positive predictive effect. This suggests that the likelihood of DWI-positive lesions in ischemic stroke patients with concomitant atrial fibrillation is higher than those without, although it does not necessarily imply a higher likelihood of DWI-positive lesions in stroke patients related to cardioembolism. A history of hyperlipidemia is the second most important positive predictive factor for DWI positivity. OMT and ONT are the third and fourth most important clinical features, respectively. As ONT and OMT duration increases, the likelihood of DWI positivity also increases; conversely, as ONT and OMT duration decreases, the likelihood of DWI negativity rises. In the LR model, the newly introduced indicator NIHSS-d(0-24h)PT is also associated with DWI presentation, serving as a positive predictive factor for DWI negativity. A larger difference in NIHSS score within 24 hours post-thrombolysis indicates a more effective thrombolysis or faster recovery, leading to a higher likelihood of DWI negativity. We observed that patients with RIS are more likely to exhibit DWI-negative results. However, the absolute value of the SHAP score for recurrent ischemic stroke is 0.19, indicating a relatively weak contribution to the LR model and suggesting a lack of strong statistical significance (Table 3; Figure 2.B).

**Figure 2.**
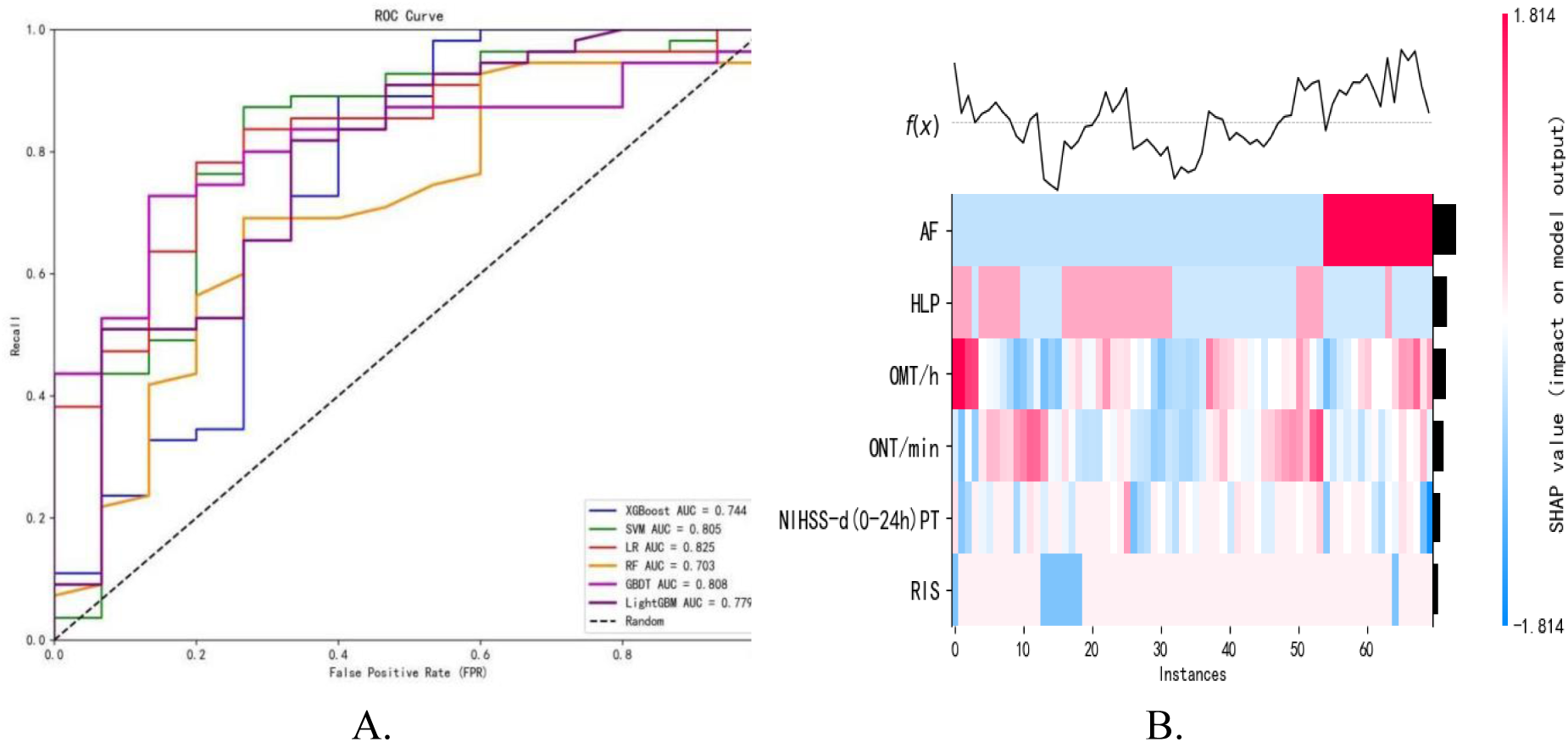
(A) ROC Curves of Six Machine Learning Models; (B) Predictive Results of Clinical Features in the Logistic Regression Model. (A) By constructing six machine learning models, including eXtreme Gradient Boosting, Support Vector Machine, Logistic Regression, Random Forest, Gradient Boosting Decision Tree, and Light Gradient Boosting Machine, the DWI outcomes were predicted. The results indicated that the Logistic Regression model performed the best, with an AUC value of 0.825, a precision of 0.930 on the test set, and a recall of 0.727. (B) SHAP values was used to interpret the Logistic Regression model, identifying the most important features in descending order of importance: AF, HLP, ONT, OMT, NIHSS-d (0-24h)PT, and RIS. Abbreviations: AF, Atrial fibrillation; HLP, Hyperlipidemia; OMT, onset to first MRI time; ONT, onset to needle time; NIHSS-d(0-24h)PT, NIHSS difference within 24 hours post-thrombolysis; RIS, Recurrent ischemic stroke.

**Table 3.**
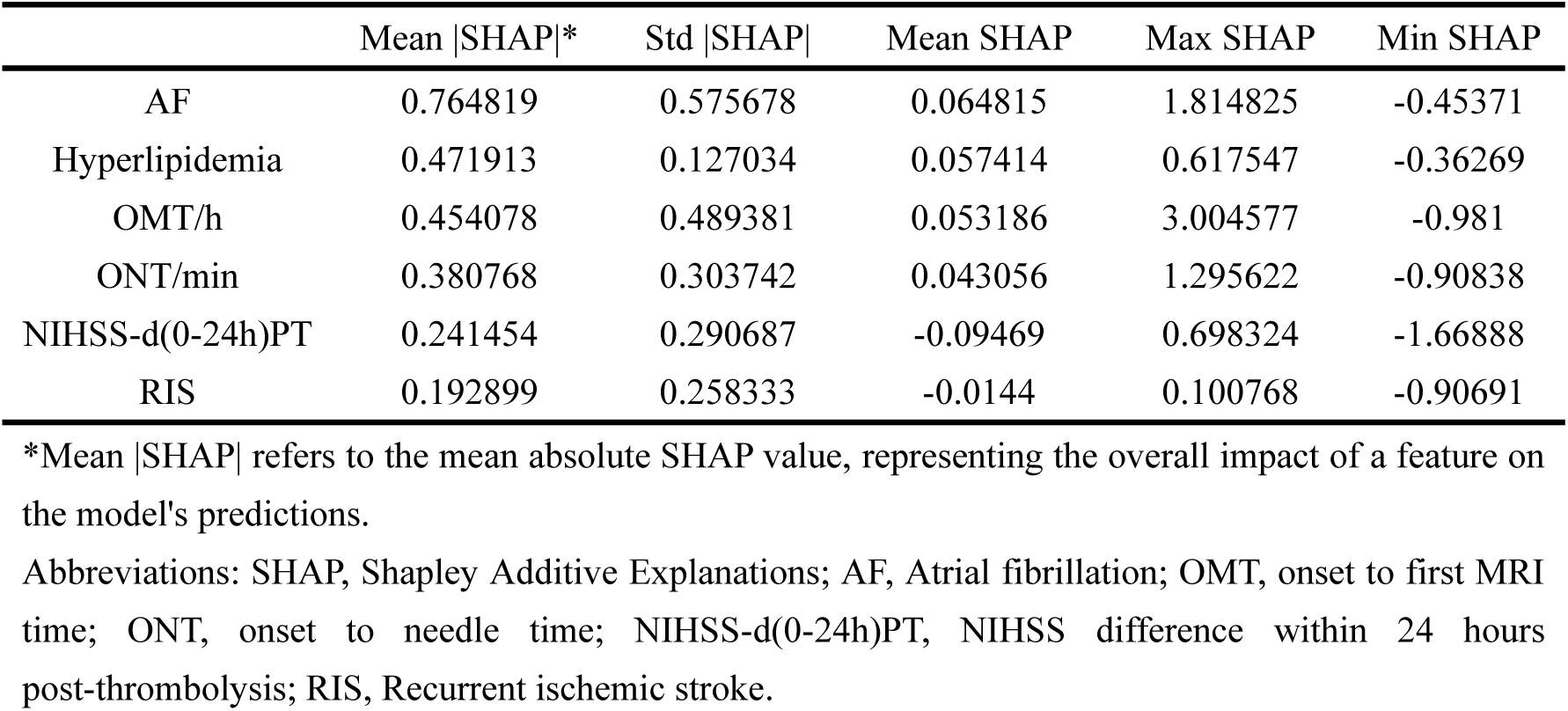
Summary of SHAP Values for the Machine Learning Model.

### 3.3 Survival Analysis

Interestingly, during telephone follow-up of DWI-negative ischemic stroke patients, we found that as of November 2023, all stroke recurrence patients had not discontinued their medication before the recurrence of symptoms. Conversely, stroke patients who self-discontinued medication after discharge never experienced a recurrence. Within one month post-stroke, three patients experienced a recurrence. Between 3 months and 1 year post-stroke, one patient experienced a recurrence, with recurrence times occurring at 4 months and 7 months after the initial DWI-negative ischemic stroke, leading to the patient’s demise due to rectal cancer one year after the stroke. One year after the stroke, two patients experienced recurrences, with one experiencing subsequent strokes at 4 years and 4.5 years post-initial stroke, and the other experiencing a recurrence 2 years later. This perplexing discovery raises questions about the risk factors for stroke recurrence in DWI-negative ischemic stroke patients. In this study, there were 63 DWI-negative ischemic stroke patients, among whom 3 died due to severe stroke, 6 passed away from conditions including cancer, myocardial infarction, heart failure, and Coronavirus Disease 2019, and one patient who succumbed to cancer experienced two stroke recurrence events before their demise. To investigate the risk of stroke recurrence in DWI-negative ischemic stroke patients, we further refined the Fine-Gray competing risk model. For DWI-negative ischemic stroke patients, over time, the cumulative incidence rates of stroke recurrence and death events both increase. However, the cumulative incidence rate of recurrent events surpasses that of death events, particularly within the first 20 months post-stroke, where the risk of recurrence far exceeds the risk of death (Figure 3). We found that age (p=0.63) and gender (p=0.61) did not exhibit statistically significant differences in their impact on recurrence risk. Compared to large artery atherosclerosis subtype, the effects of small artery occlusion (p=0.33) and cardioembolic stroke (p=0.38) on stroke recurrence risk were not statistically significant. However, stroke patients of other Determined Etiology (p=0.00) and Undetermined Etiology (p=0.00) showed a significantly reduced risk of recurrence (Table 4). Given the limited number of DWI-negative ischemic stroke patients in this study, the statistical significance of the results may be influenced. Further validation of the above findings will require larger sample sizes and longer follow-up periods in prospective studies. Future research could delve deeper into the risk factors for stroke recurrence in DWI-negative patients, the relationship between stroke etiology types and recurrence, as well as the association between stroke recurrence outcomes and medication adherence. This study was constrained by the small dataset and did not address the aforementioned aspects.

**Figure 3.**
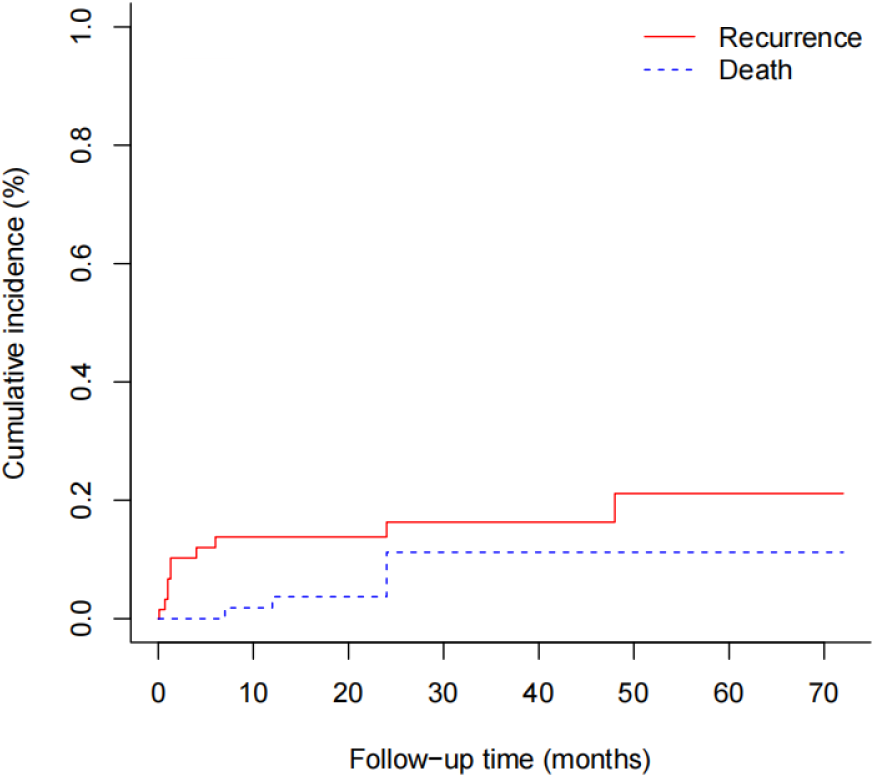
Cumulative Incidence of Stroke Recurrence and Death. Using the Fine-Gray competing risk model, we analyzed the risk of recurrence and death in DWI-negative ischemic stroke patients. The x-axis represents the follow-up time after DWI-negative ischemic stroke, while the y-axis represents the cumulative incidence risk of events. The solid red line indicates the cumulative risk of stroke recurrence, and the dashed blue line represents the cumulative risk of death after stroke.

**Table 4.** The impact of clinical characteristics on the risk of recurrence in the Fine-Gray competing risk model.

|  | HR | CI 95%_lower | CI 95%_upper | p |
| --- | --- | --- | --- | --- |
| Age | 1.01 | 0.14 | 7.40 | 0.63 |
| Sex | 1.43 | 0.09 | 23.56 | 0.61 |
| SAO vs LAA | 0.49 | 0.19 | 1.29 | 0.33 |
| CE vs LAA | 2.08 | 0.04 | 122.66 | 0.38 |
| SOE vs LAA | $3.14 \times 10^{-5}$ | $3.14 \times 10^{-5}$ | $3.14 \times 10^{-5}$ | 0.00 |
| SUE vs LAA | $4.60 \times 10^{-5}$ | $4.60 \times 10^{-5}$ | $4.60 \times 10^{-5}$ | 0.00 |

Abbreviations: LAA, Large-artery Atherosclerosis; CE, Cardioembolism; SAO, Small-artery Occlusion; SOE, Stroke of Other Determined Etiology; SUE, Stroke of Undetermined Etiology.

## 4. Disccusion

DWI is considered a crucial diagnostic tool for cerebral infarction, yet a subset of ischemic stroke patients present as DWI-negative on imaging. The aim of this study is not to negate the value of DWI scans in diagnosing stroke but to provide insights into DWI-negative ischemic stroke, aiming to enhance accurate diagnosis, reduce misdiagnosis and underdiagnosis of DWI-negative stroke, and provide prognostic information for DWI-negative stroke. In this study, we analyzed the imaging characteristics and relevant clinical features of acute ischemic stroke patients diagnosed after intravenous thrombolysis. We found that multiple factors influence the imaging presentation on cranial magnetic resonance imaging, including NIHSS-1w, TOAST, OMT, ONT, PLT, RIS, AF, and HLP (Table 2). In this study, there were no statistically significant differences in age and gender between the DWI-positive and negative groups, consistent with previous research findings[11, 12]. However, the two groups exhibited significant differences in NIHSS scores. Following adjustment for confounding factors in binary logistic analysis, we identified the NIHSS-1w as the most critical factor. Furthermore, DWI-negative ischemic stroke patients consistently had lower NIHSS scores at four time points compared to DWI-positive patients, suggesting that most DWI-negative stroke patients had minimal neurological deficits after intravenous thrombolysis treatment, with better recovery at 1 week of hospitalization, indicative of mild stroke, aligning with previous research conclusions[13]. The TOAST classification also showed significant statistical differences between the two groups. The DWI-negative group predominantly consisted of small artery occlusion strokes (67.7%), while the DWI-positive group mainly had large artery atherosclerosis strokes (39.7%). We believe that lacunar strokes are more likely to present as DWI-negative after thrombolysis, consistent with previous research findings[14]. This may be attributed to a higher likelihood of complete reperfusion in small vessel disease after thrombolysis, while partial reperfusion may be more common in large vessel disease. However, the specific impact of stroke etiology types on DWI presentation remains unclear, possibly due to the small sample size in the DWI-negative group. There were significant statistical differences between the two groups in important time intervals during the course of cerebral infarction. OMT and ONT remained statistically significant after adjusting for confounding factors and showed statistical differences in the LR model. In this study, DWI-negative ischemic stroke patients had a significantly longer time to complete the initial DWI scan (89.33±49.65 hours) compared to the limited time for lesion identification with DWI sequences[15]. The median time from symptom onset to thrombolysis was around 2 hours. We cannot use the detection time window of DWI sequences in acute cerebral infarction as a reference indicator for diagnosing DWI-negative stroke. We believe that the reason for DWI negativity is that thrombolysis treatment takes precedence over DWI scans, allowing for rapid salvage of the ischemic penumbra. Prior research also supports that earlier thrombolysis leads to shorter ONT, timely reperfusion of the ischemic area, and a higher likelihood of DWI signal reversal from high to normal[16, 17]. Furthermore, our study found that the medical history and comorbidities of stroke patients also have predictive value for DWI imaging characteristics. Specifically, there were statistically significant differences between the two groups in terms of AF, HLP, and RIS. Among these, AF had the highest predictive value and indicated a higher incidence of DWI positivity, consistent with some previous research conclusions[9]. This may be because AF often suggests cardioembolic stroke, which tends to be a more extensive and severe type of stroke[18]. However, it is important to note that not all stroke patients with concomitant AF have cardioembolic strokes. Furthermore, patients with HLP were more likely to have DWI-positive strokes, possibly related to the propensity of hyperlipidemia patients to experience anterior circulation and larger area cerebral infarctions[19]. The specific underlying mechanisms remain unclear and warrant further investigation. Lastly, patients with a history of stroke are more likely to have DWI-negative recurrent strokes. Some studies suggest that prior cerebral infarctions may promote the development of more collateral circulation in the brain, slowing the progression of the ischemic penumbra towards the infarcted area during recurrent strokes[20]. However, the statistical significance level between the two groups may be subject to the “edge significance” trap, and its predictive value is relatively low (Mean |SHAP|: 0.19). Future large-scale, multicenter studies are needed to further explore this relationship. Some studies suggest that higher platelet counts may increase the risk of large vessel thrombosis, coagulation abnormalities, and vasculitis, leading to multiple ischemic lesions manifesting as high signal intensity on DWI[21, 22]. However, we found that an increase in PLT was associated with a higher probability of DWI-negative strokes, although the correlation was modest, necessitating further validation with a larger sample size. Additionally, we observed that for patients with DWI-negative ischemic stroke, the risk of recurrence in the short term outweighs the risk of death, and the TOAST classification may be related to the risk of stroke recurrence. However, due to sample size limitations, the evidence supporting this relationship remains insufficient. In conclusion, previous studies have identified four main reasons for DWI-negative strokes in patients with cerebral infarction: misdiagnosis, posterior circulation infarction, small infarct volume, and rapid completion of DWI scans before thrombolysis treatment[2, 8]. Our study suggests that patients with mild stroke, small infarct volume, rapid intravenous thrombolysis treatment, and recurrent ischemic stroke tend to present as DWI-negative strokes.

Although our study has clinical significance, it has certain limitations. Firstly, the sample size of DWI-negative strokes in this study is relatively small compared to DWI-positive strokes, which may impact the statistical power of the analysis and the reliability of the results. Therefore, we have interpreted the results by combining odds ratios and confidence intervals, and future clinical studies should aim to collect a larger sample of DWI-negative patients. Secondly, there were missing clinical data for some patients in this study, and we addressed this by simple imputation, which may introduce bias and increase the inaccuracy of the results. Thirdly, while this study included follow-up of DWI-negative stroke patients, the duration of patient follow-up varied, making it challenging to conduct robust survival analysis. Future research should strengthen follow-up of DWI-negative patients, increase the sample size, and further explore risk factors for adverse outcomes, including medication adherence, TOAST classification, and their impact on the recurrence of DWI-negative strokes.

## 5. Conclusion

For acute ischemic stroke patients undergoing intravenous thrombolysis, the incidence of DWI-negative strokes is 22.74%. Factors positively associated with DWI-positive lesions include NIHSS-1w, ONT, OMT, HLP, and AF. Conversely, these factors serve as negative predictors for DWI-negative strokes. PLT and RIS are negative independent predictors for DWI-positive lesions, while they act as positive predictors for DWI-negative strokes. The predictive value of AF and HLP is the highest, while the predictive value of PLT and RIS is relatively low. In patients with DWI-negative ischemic stroke, the risk of recurrence in the short term outweighs the risk of death post-onset, and the TOAST classification may be associated with the risk of stroke recurrence, highlighting the need for enhanced follow-up and secondary prevention measures.

## Data Availability

All data presented in this manuscript are genuine and verified

## Acknowledgments

We would like to thank Cheng Huang for providing the patient list from the stroke center, Yangtao Liu and Yingli Li for their guidance on statistical analysis, and Jun Mu for advising on manuscript writing and revisions. We also extend our gratitude to Jie Zhang, Mi Xiao, Zhiwen Yan, and Huan Zhao for their contributions to drafting parts of the manuscript.

## Non-standard Abbreviations and Acronyms

DWI: Diffusion Weighted Imaging
TOAST: Trial of Org 10172 in Acute Stroke Treatment
ODT: onset to hospital-door time
DNT: door to needle time
ONT: onset to needle time
OMT: onset to first MRI time
OHDT: onset to hospital discharge time
NIHSS-iPT: NIHSS score immediately post-thrombolysis
NIHSS-24hPT: NIHSS score at 24 hours post-thrombolysis
NIHSS-1w: NIHSS score at 1 week of hospitalization
NIHSS-HD: NIHSS score at hospital discharge
NIHSS-d(0-24h)PT: NIHSS difference within 24 hours post-thrombolysis
HLP: Hyperlipidemia
AF: Atrial fibrillation
TIA: Transient ischemic attack
RIS: Recurrent ischemic stroke
CHD: Coronary heart disease
CRP: C-reactive protein
Hb: Hemoglobin
PLT,: Platelet
D-D: D-dimers
Tc: Total Cholesterol
LDL-C: Low-Density Lipoprotein Cholesterol
HbA1c: Hemoglobin A1c
UA: Uric acid
LAA: Large-artery Atherosclerosis
CE: Cardioembolism
SAO: Small-artery Occlusion
SOE: Stroke of Other Determined Etiology
SUE: Stroke of Undetermined Etiology
SVC: Support Vector Classification
LR: Logistic Regression
AUC: Area Under the Curve
ROC: Receiver Operating Characteristic

